# Trends and Variation in Data Quality on the EU Clinical Trials Register: A Cross-Sectional Study

**DOI:** 10.1101/2021.06.29.21259627

**Authors:** Nicholas J. DeVito, Ben Goldacre

## Abstract

The EU Clinical Trial Register (EUCTR) is a public facing portal containing information on trials of medicinal products conducted in the European Union (EU) and European Economic Area (EEA). Today, the registry holds information on over 30,000 trials. Given its distinct regulatory purpose, and results reporting requirements, the EUCTR should be a valuable open-source hub for trial information. Past work examining the EUCTR has suggested that data quality on the registry may be lacking. Using the full EUCTR public dataset, we examined areas in which national regulators are expected to ensure data quality including the posting of registrations, updating trial completion information, and monitoring results posting in line with EU guidelines. We identified issues across all areas examined with notable research hubs like France, Spain, and The Netherlands lacking consistent and complete data on the registry. These deficiencies complicate the utility of the EUCTR for research, transparency, and accountability efforts.

## Introduction

Registration of clinical trials in a publicly accessible registry is a key tool in ensuring transparency and accountability in clinical research. Prospective and sufficiently detailed registrations allow public accounting of planned, ongoing, and completed research, provides information on trial design and outcomes, and shares this data in a convenient and accessible format. This is vital information for clinicians, researchers, health officials and the public. Registries should provide a single, open source, repository for key information on clinical research. These benefits can only be realised, however, when a registry is functioning as intended.

The EU Clinical Trials Register (EUCTR), and its back-end EudraCT system, acts as a World Health Organisation International Clinical Trial Reporting Platform (ICTRP) primary registry.^1^ This means registration data from the EUCTR flows to the ICTRP master dataset of global clinical trials, and prospective registration on the EUCTR would meet the requirements for trial publication imposed by a number of leading academic journals.^2^ It also serves as the official regulatory registry for interventional clinical trials of investigational medicinal products (CTIMPS) in the European Union (EU) and European Economic Area (EEA). All CTIMPs in the EU and EEA should be registered in the EudraCT system and most should then appear on the public EUCTR. Phase 1 trials in healthy adults are exempt from public posting. CTIMP sponsors are required to file a protocol with the national competent authority (NCA) in each EU/EEA country with planned enrollment for that trial. This information is then signed off on by the regulator and sent to the EUCTR which is managed by the European Medicines Agency (EMA).^3^ Once the appropriate ethics and regulatory approvals are in place, a country-specific protocol for each EU/EEA location, referred to as a Clinical Trials Application (CTA), is posted to the public EUCTR record for that trial.^4^ EU guidelines also require sponsors of all registered trials to post results to the registry.^5^ Since 2015 these must be posted in a standardised tabular format within a year of the completion of the trial, and within six months for paediatric trials.^5,6^

Prior work has suggested that several issues may compromise the EUCTR’s utility as a valuable source of information about European clinical trials. For example trial status data on the EUCTR is often outdated compared to the same trial registered on ClinicalTrials.gov.^7^ The EU TrialsTracker project tracks reporting of trials on the EUCTR as required under EU guidelines^8^ and relies on accurate data on trial completion to function. As of December 2020 there were are 35,000 unique registrations on the EUCTR and 8,955 of 13,152 (68.1%) verifiably due trials have reported; however over 8,000 trials cannot be properly assessed due to data inconsistencies, and even more appear categorized as “Ongoing” when they likely completed long ago. Delays in setting up links to NCAs and implementing a data verification system has led to known data issues such as missing completion or trial status data for records up to March 2011 (i.e., “historical data”).^9^ According to the EUCTR website, the EMA is working with NCAs “to ensure key data on the status of existing trials is complete.”^9,10^ Progress on this front, however, is not documented and appears inconsistent.

Given the size and clinical research output of the EU/EEA, the EUCTR should provide a wealth of public information about clinical trials and promote accountability in their reporting. However issues with data quality and completeness may compromise this functionality. As there is no readily available information or documentation as to the extent of potential data issues, we set out to examine and describe trends in NCA-level registration and reporting practices on the EUCTR.

## Methods

### Data Collection

We used scraping software to collect data from each public country-level protocol on the EUCTR (i.e., all CTAs) as of 1 December 2020. This was the last month in which full UK data was available on the EUCTR prior to leaving the EU and could therefore be compared to its European peers. As of 1 January 2021 UK sponsors may still add results to the EUCTR for existing registrations but protocol adjustments, including updates to trial status and completion dates, are not possible and any ongoing CTAs are tagged as no longer under the purview of the EMA.^11^ Box 1 contains a typical trial record on the EUCTR.

#### Box 1

Example of an EUCTR Trial Record

*An EUCTR trial record contains links to all country-level CTAs in the “Trial protocol” field, and a link to results, if available, in the “Trial results” field. The individual country CTAs contain detailed information on the trial including the date the NCA entered the record into the registry and completion information. The results section can contain information on enrollment and a clearer “Start Date” value than the one provided in the upper-right of the trial record which is not tied to enrollment but rather ethics and regulatory approval*.

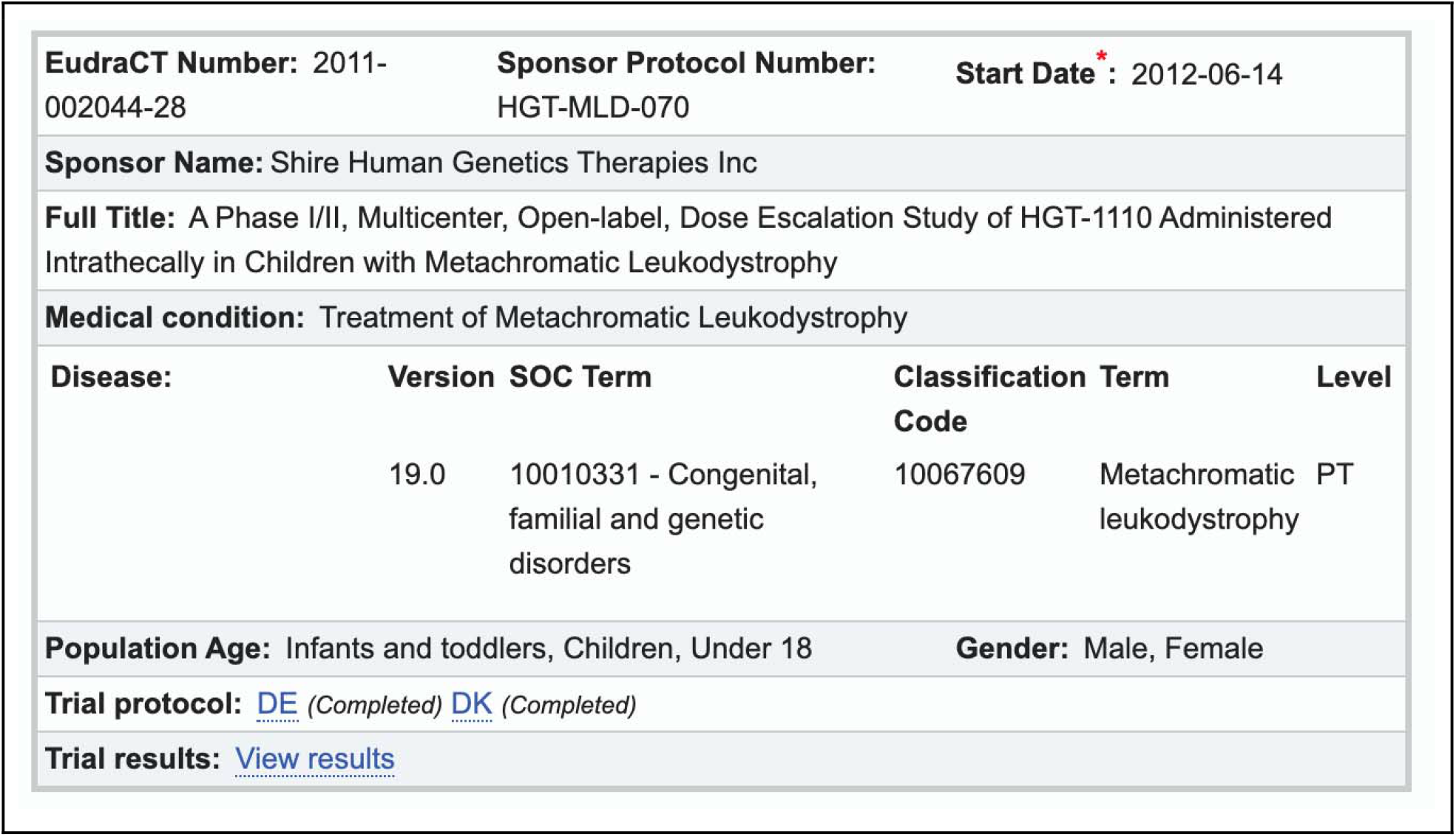

### Study Population

All EU trial records linked to an EU/EEA country as of December 2020 were included in our analysis. Certain paediatric trials include non-EU/EEA CTAs and these were excluded as they are not linked to any individual NCA and lack detailed information on trial completion by design. The relevant NCA for a given CTA was identified by the “National Competent Authority” field in the EUCTR protocol “Summary” section. Germany has two independent NCAs that manage trial records and these were examined separately throughout unless otherwise noted. Information on NCAs for all EU/EEA countries as of December 2020 (per the EudraCT website) are available in Table 1.^12^

**Table 1.**
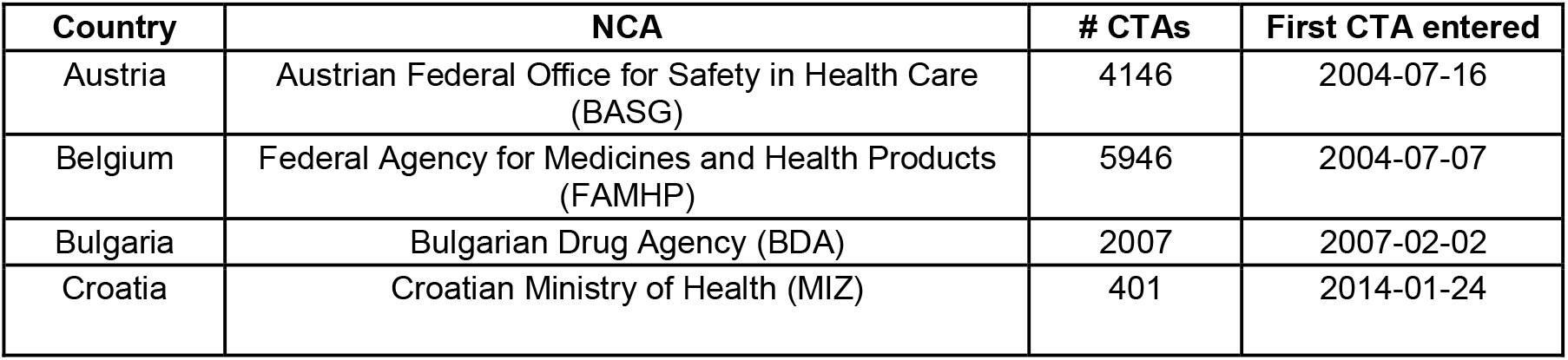
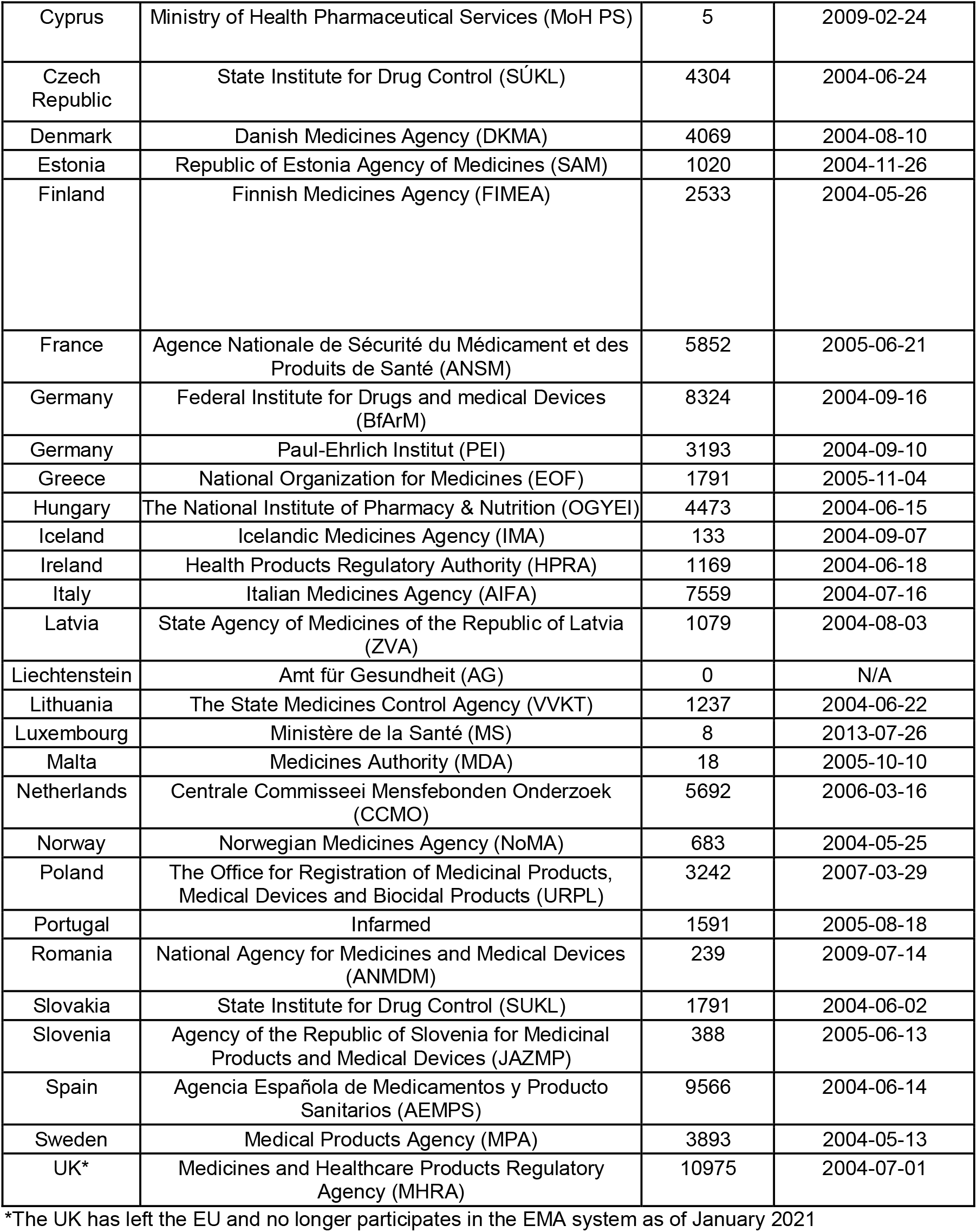
Details of EU/EEA National Competent Authorities (NCAs):

### Trends in CTIMP Registration by NCA

We describe the trend in new registrations on the EUCTR by NCA over time. Each EUCTR CTA contains a field denoting the “Date on which this record was first entered in the EudraCT database” (i.e., the record entry date). While not necessarily indicative of when information was first submitted to the NCA (this information is not available in the EUCTR), this date represents when the NCA first entered the protocol information into the EudraCT system and so should act as a proxy for NCA, rather than sponsor, registration activity.^13^ Trends for EUCTR entry date were compared to trends for NCA approval dates as a check for consistency. We show the overall trend in new CTA registrations and unique trials for each full year in the dataset (2005-2019) and the cumulative number of new CTAs for each NCA.

Prior experience from the UK has shown that administrative issues can cause delays or issues in registrations appearing on the public EUCTR website.^14,15^ In order to examine whether missing CTAs is an issue in other EU/EEA countries, we selected all trials in the database that had results available in the EUCTR’s tabular format. This format includes a standard data field indicating which countries enrolled participants in the trial. Using a custom web scraping program, we extracted all enrollment countries from each trial and compared them to the CTAs associated with the trial registration. In practice, every EU/EEA location with confirmed enrollment in the results should have a public CTA associated with that trial. However, some trials may include current EU/EEA locations prior to either their entry into the EU/EEA or their linkage to the EMA regulatory system, and therefore would not have a CTA; as such we only expected an enrollment country to have an associated CTA when the trial start date (also taken from the tabular results) was later than the earliest known CTA on the registry from that NCA (see Table 1). We report the expected vs. actual CTAs based on the results information for each country and over time. For the time trend, we report the total number of CTAs that were expected but could not be located, and the trend in missing CTAs as a percent of all public CTAs entered in a given year.

### Quality of Trial Status and Completion Date Fields

The current status of clinical trials in each country should be clear from the “Trial Status” field, and eventually the “Date of the Global End of the Trial” field. These should indicate, in each CTA, when the trial has completed in all countries.^3^ Trial completion information is also available in the results section, but this is only available for trials that have results and it is not linked to official end of trial paperwork filed with an NCA. We show the distribution of CTA trial status’ on the EUCTR overall and broken down by the responsible NCA. Then, limiting the population only to CTAs that are in a “Completed” or “Prematurely ended” status, we examined the availability of the “date of the global end of the trial” field and distribution over time by NCA.

### Results Availability

EU Guidelines (Section 4.7) call on member states to “verify that for clinical trials authorised by them the result-related information is posted to the Agency” and non-reporting after 15 months “will be flagged…[and] publicly available.”^5^ While the EUCTR does not currently include any official flags for non-reporting, the availability of results over time can be assessed independent of completion status or dates to identify irregular trends. We separated all trials in our population into those that have a single EU/EEA CTA, and those that have multiple EU/EEA CTAs. Since reporting on the EUCTR occurs at the trial level, and not the CTA level, for trials with only a single CTA, the responsibility for reporting follow-up falls solely within the remit of that NCA. We report the proportion of all trials with results available on the EUCTR by year and for single-CTA and multi-CTA trial sub-populations. We then examined the trends in reporting for single-CTA trials as the responsibility for follow-up would sit only with the relevant NCA. Results status was examined through the presence of a “View Results” link in the Summary section of a country level protocol. This link is only available when a trial has results available.

### Data and Code Availability

Data analysis was performed in Python 3.8.1 and all data and code can be accessed on GitHub at: https://github.com/ebmdatalab/euctr_data_quality. EUCTR scrapers can be accessed at: https://github.com/ebmdatalab/euctr-tracker-code and https://github.com/ebmdatalab/registry_scrapers_parsers

## Results

As of 1 December 2020, the EUCTR contained 98,622 CTAs across 38,566 registered trials since 2004. Removing all non-EU/EEA CTAs leaves 97,227 CTAs across 37,520 trials. Table 1, above, shows the total number of registered protocols from each NCA and the earliest entered CTA for a given NCA on the registry. Non-EU/EEA CTAs were excluded from all analyses.

### Trends in Registration on the EUCTR

Figure 1 shows the overall trend in new CTA registrations and overall trials by year of first CTA entry.

**Figure 1:**
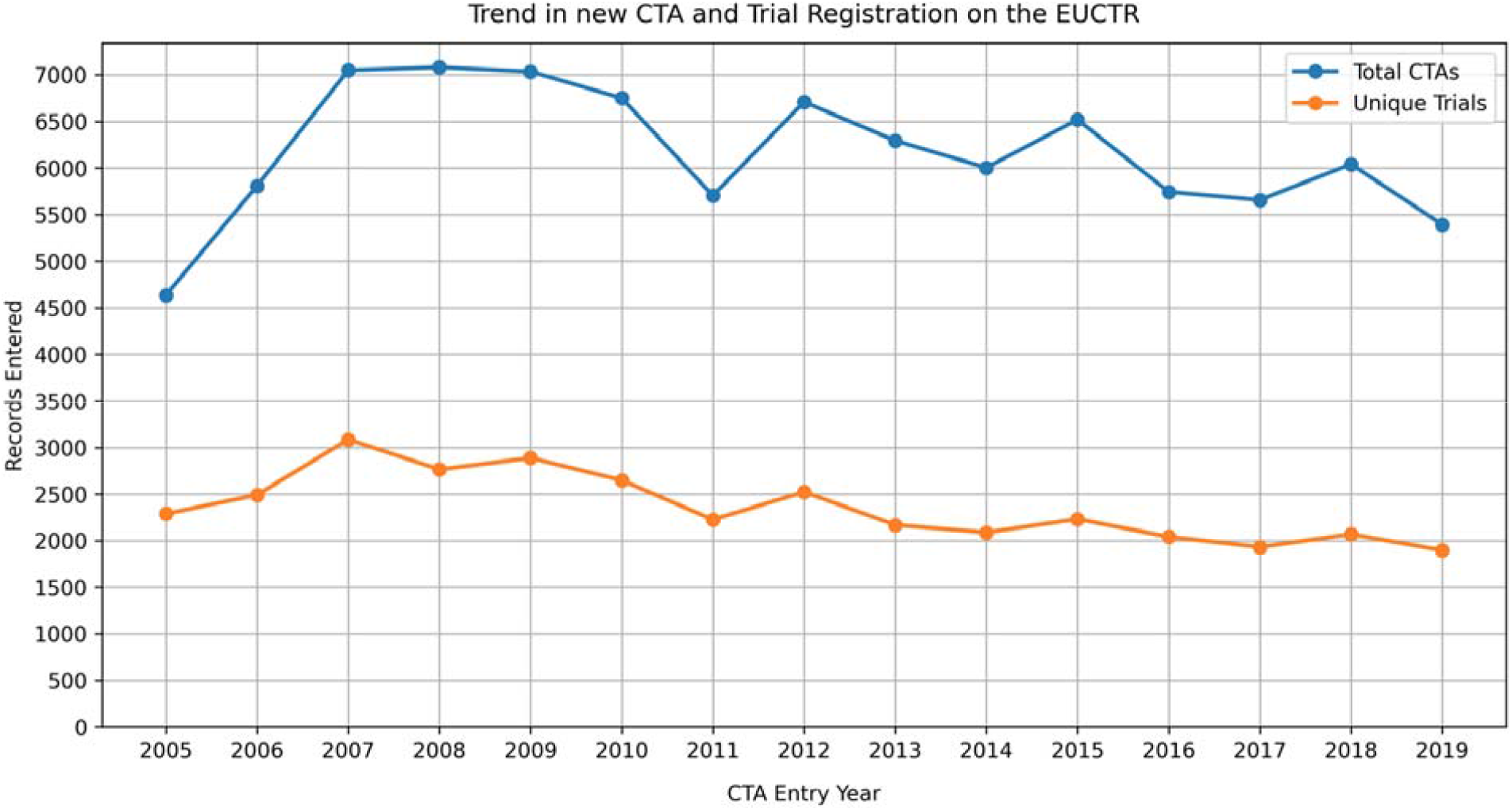
The overall trend in number of EU/EEA trials, and in total CTAs for all years with full data (i.e., excluding 2004 and 2020). A single trial registration on the EUCTR is made up of individual EU/EEA CTAs for countries with current or planned recruitment.

Figure 2 shows the trend in cumulative number of CTAs for each NCA. NCAs with under 100 registered CTAs (Cyprus (n=5), Luxembourg (n=8), Malta (n=18), and Liechtenstein (n=0)) are excluded from NCA trend breakdowns throughout. A graph of new registrations by quarter is available in Supplemental Figure 1. The annual trend in new registrations closely matched trends in NCA approval dates so results would now change substantially based on the usage of either date for the analysis (Supplemental Figure 2).

**Figure 2:**
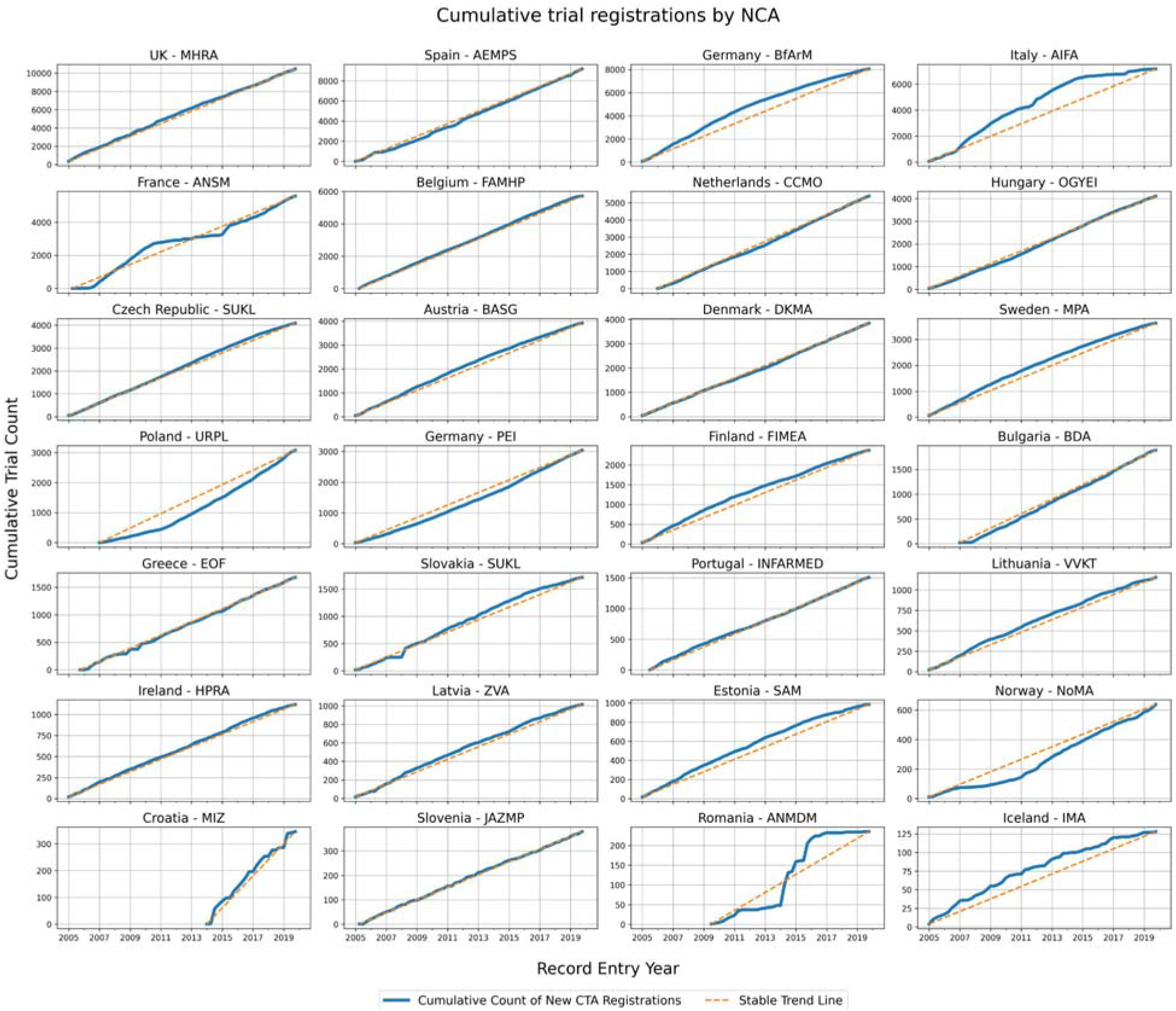
The cumulative number of new trials for each EU/EEA country from 2004 through 1 December 2020. Data was aggregated by quarter. NCAs are listed in order of total CTAs. A stable trend line, assuming the same number of registrations are added every quarter, is shown in orange for each country to aid in visualising trends in new CTA registrations over time. NCAs are ordered by total number of registered CTAs.

Most NCAs register new trials at a steady rate with minimal variation over time, with some showing slightly decreasing trends in new-CTAs (e.g., Estonia, Finland, Sweden, Iceland). Other countries had more notable trends or distinct abnormalities. France had a plateau in new CTAs between 2010 and 2015 and Italy experienced a prolonged slowing of new CTAs eventually leading to a plateau in 2015. Romania shows 2 separate plateaus for new trials between 2011 and 2014 and again from 2016 through the present with a relative surge in new CTAs in between. Germany’s two NCAs displayed opposing trends with new BfArM CTAs decreasing and new PEI CTAs increasing over time. Both Poland and Norway showed an acceleration in new CTAs over time.

Figure 3 shows how often the countries reported in the results section matched the CTAs associated with that trial: 22 of the 30 (73%) EU/EEA countries had more than 90% of expected protocols available. Only Croatia had all expected protocols publicly available. Consistent with the outlier trends data discussed above, France, Italy, Poland, Norway, and Romania all appeared to be missing substantial numbers of expected CTAs compared to peer countries. France, Norway, and Romania displayed particularly low CTA availability with under 50% of expected CTAs available in the public dataset. Although their expected trial count was very low (n=6), all Cypriot trials were missing. Figure 4 shows the trend in missing CTAs, and as a percent of all publicly available CTAs, by year. This confirms that the issue is not confined to older or “historic” data or NCA linkage issues as high levels of missing CTAs persist from 2010-2016 with a tail-off only occurring in recent years where most results would not yet be available to check for matching CTAs.

**Figure 3:**
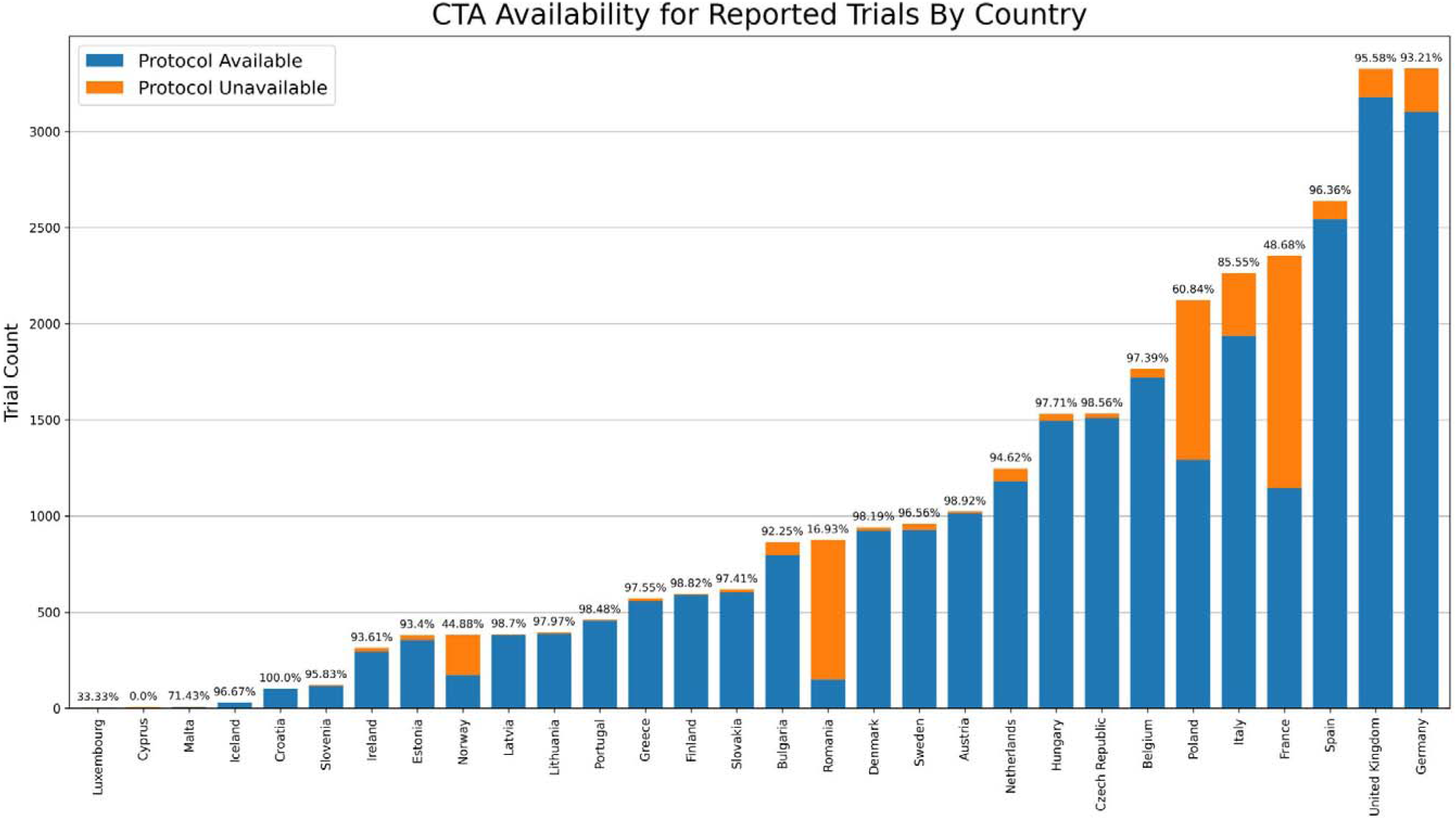
The number, and percent, of CTAs that should be publicly available for all trials with tabular results on the EUCTR. The CTAs available for each country were compared to detailed tabular results, where available, that contains information on which specific countries enrollment occurred.

**Figure 4:**
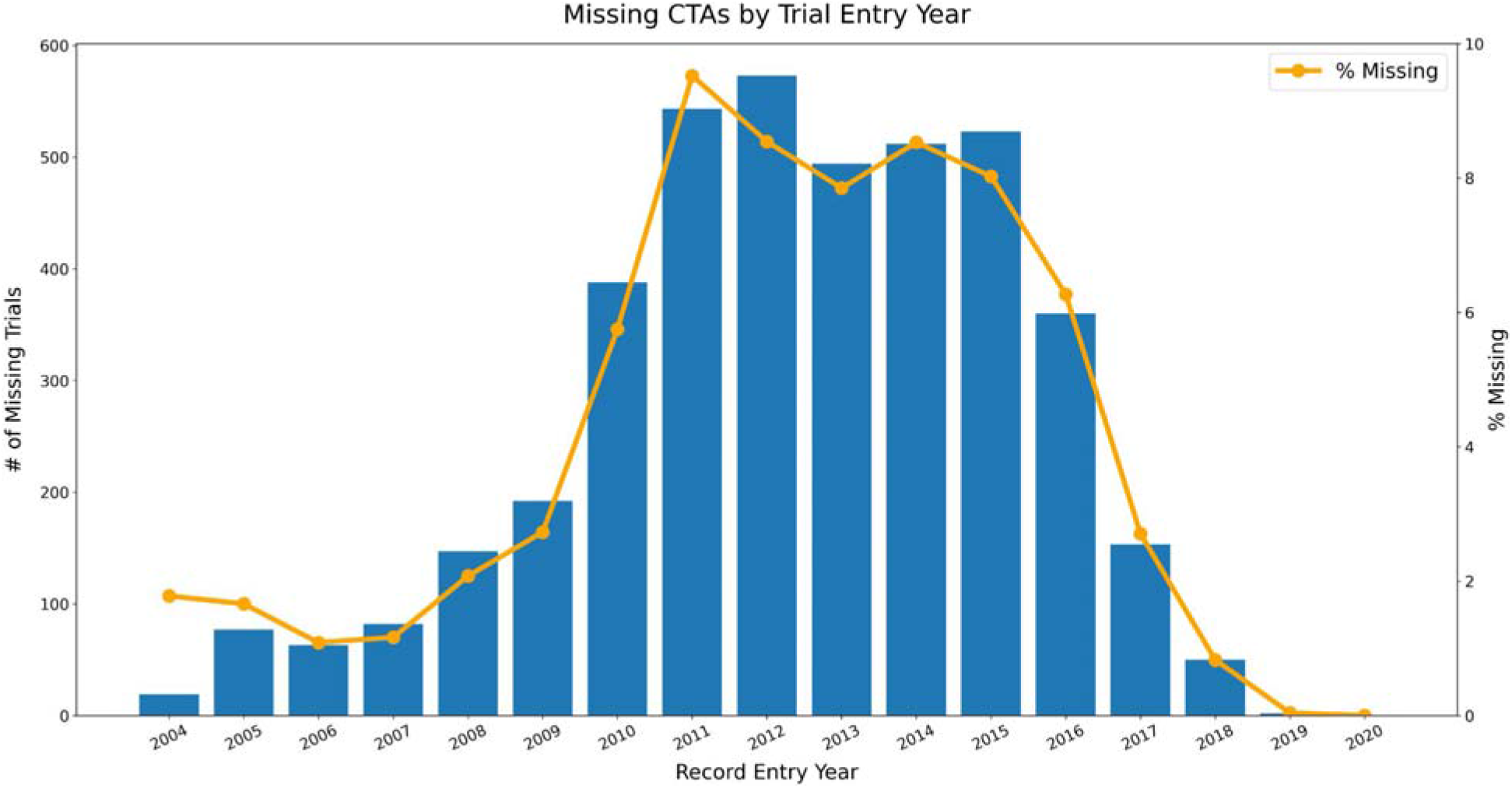
The bars represent the total number of missing CTAs by the year in which the earliest public CTA was entered for the parent trial registration. The line represents the missing CTAs for trials first entered in that year as a percentage of all public CTAs first entered in that year.

### Completion Status Trends

Figure 5 shows the number of trials in each trial status available on the EUCTR by registration year, ordered from the highest to the lowest proportion of completed CTAs. The overall trend is available in Supplemental Figure 3. Most countries display a common pattern in which the vast majority of older trials are completed with an increasing number of newer trials listed as “Ongoing”, as would be expected. Deviations from this trend were minor in some countries (e.g., Belgium, Italy, Sweden) and pronounced in others (e.g., Spain, Netherlands, Norway). The issues with missing trials in France and Romania appear compounded by high rates of “ongoing” older trials with issues not limited to the historic pre-2011 dataset.

**Figure 5:**
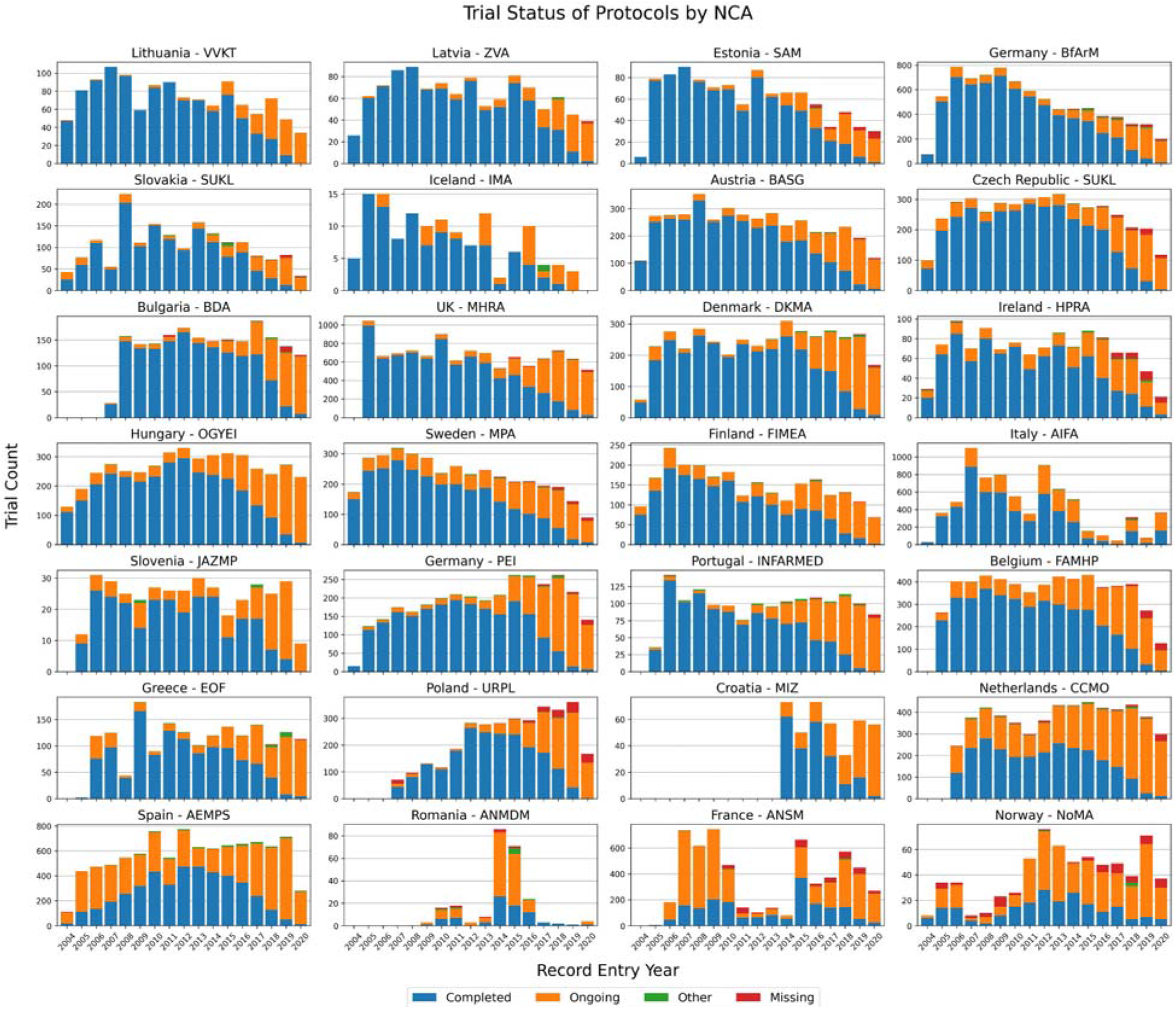
The distribution of trial status of each NCA CTA by year. NCAs are ordered by the overall percent of CTAs in a “completed” status. The expectation would be that most older trials are completed and with a slow taper for newer trials. The “completed” category includes trials in status “Completed” or “Prematurely Ended” and the “Ongoing” category also includes trials in status “Restarted.” The “Other” category includes the “Not Authorised” (n=73), “Prohibited by CA” (n=38), and “Suspended by CA” (n=22) statuses. Instances in which the trial status was missing from the “trial status” field are also noted.

Figure 6 shows the trend in CTAs in a “completed” status that have a completion date in the “Date of the Global End of Trial” field. The overall trend in completion date availability is provided in Supplemental Figure 4. This field should be updated by the NCA when a trial completes so it is clear when results are expected. Here many of the same countries with trial status issues also fail to provide completion dates for their CTAs: Spain, Italy, Belgium, and the Netherlands all have substantial protocols with missing dates beyond the 2011 cut-off for “historic” data. Germany (PEI) also appears to have a consistent level of missing dates over time. Other patterns of missing dates do appear restricted to the pre-2011 “historic” dataset (e.g., Latvia, Slovenia, Ireland).

**Figure 6:**
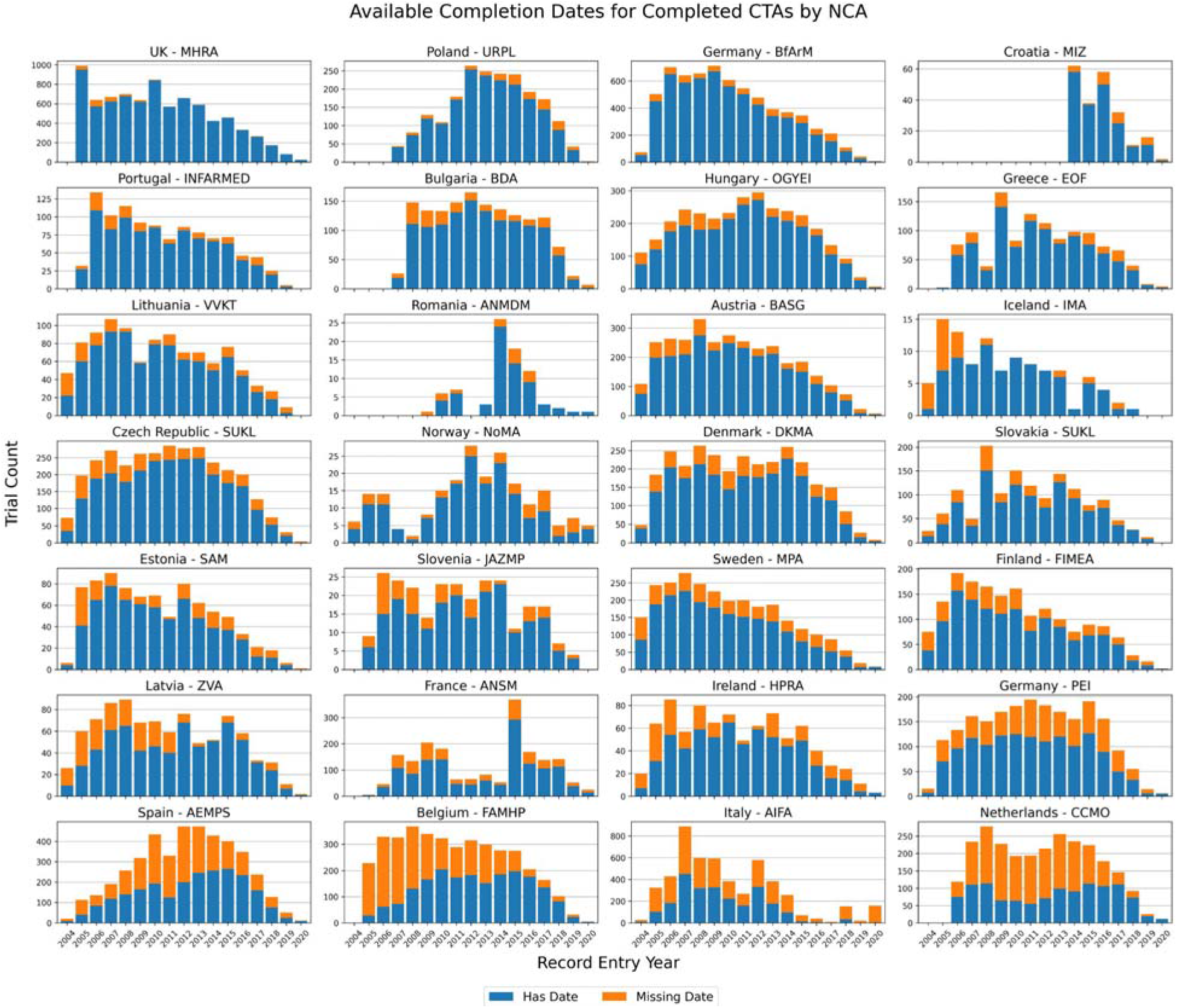
The percent of CTAs from each NCA which have a “completed” or “prematurely ended” status that also have a “date of the global end of the trial” data element, by year. While some more recent trials may not be completed globally, most older trials should ideally have a completion date on record with the NCA collected via end of trial paperwork.

### Results Availability

Figure 7 shows the trend in results availability for all registered trials (n=37,520), and split for trials with a single CTA (n=23,623) and multiple CTAs (n=13,897). Reporting is consistently and substantially lower for single EU/EEA country trials compared to trials with multiple EU/EEA CTAs.

**Figure 7:**
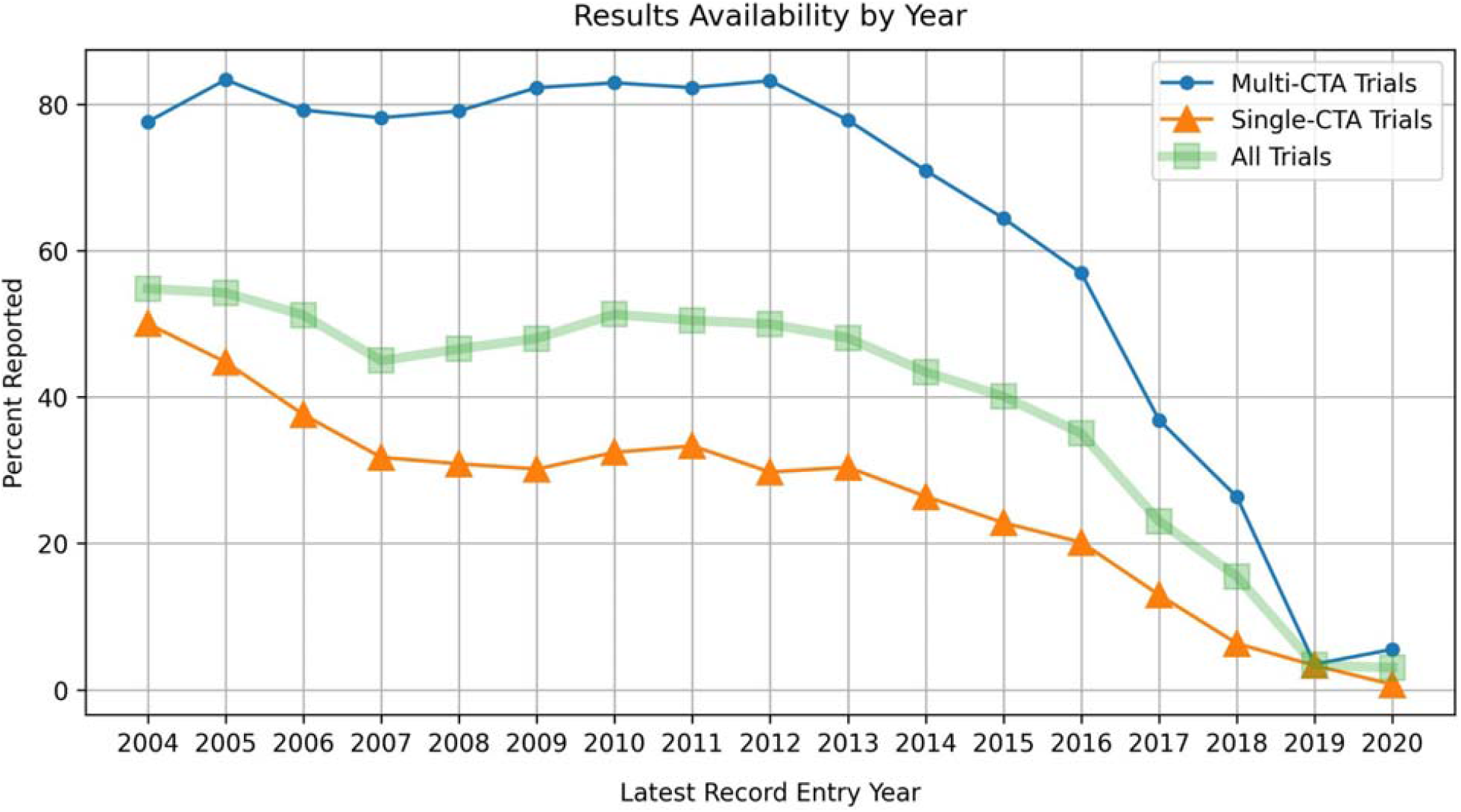
The percent of trials that have an associated results report by year of the latest CTA-record entry for a given trial overall, for trials with a single CTA and for trials with multiple EU/EEA CTAs. The drop-off in results for more recent trials is expected as many of these either will have only recently completed or are still ongoing.

Figures 8 shows the NCA-level reporting trends for single-CTA trials. Trials taking place in a single EU/EEA country were poorly reported across all NCAs with the UK representing a notable exception compared to their peer nations.

**Figure 8:**
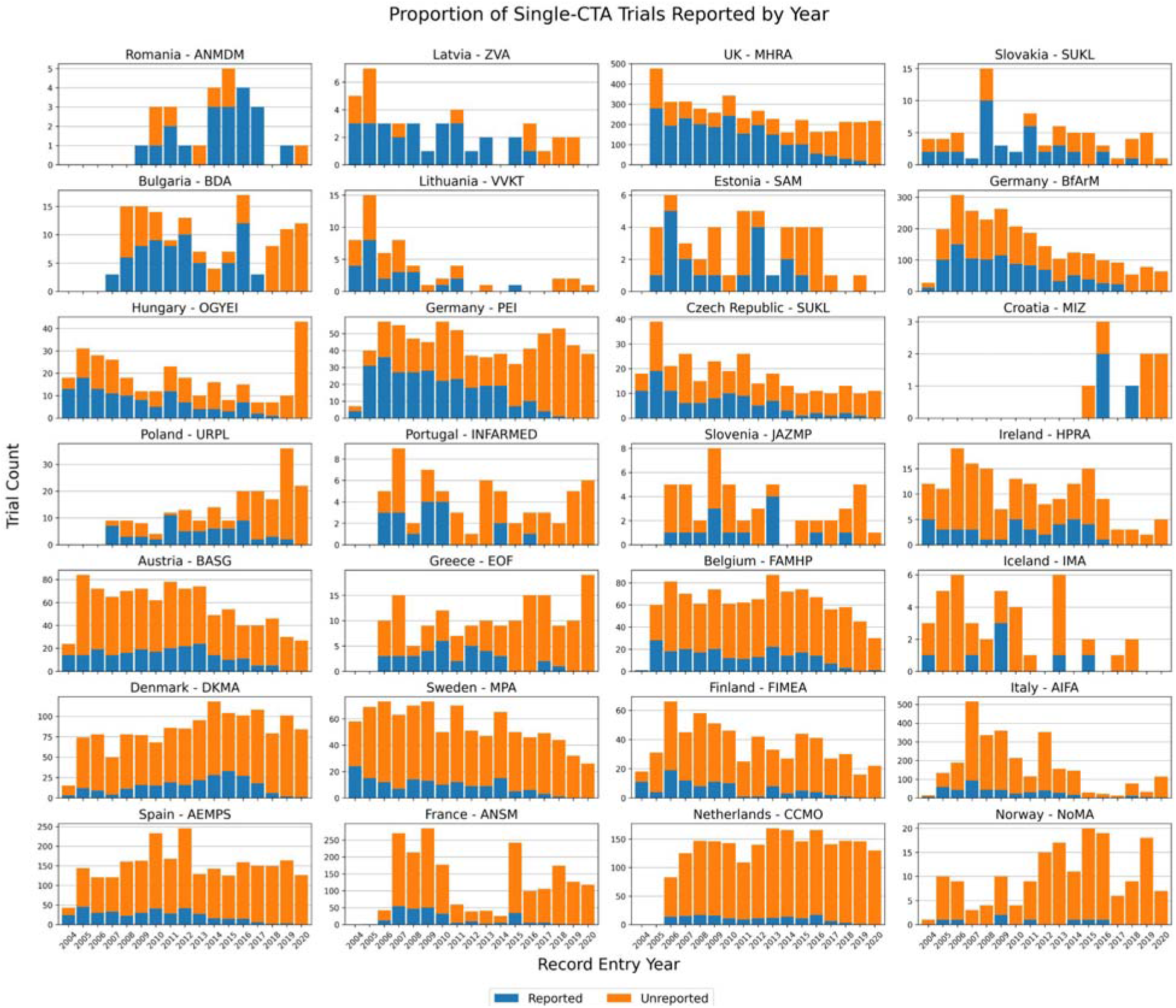
The trend in the proportion of trials, for each NCA, in which only a single EU/EEA CTA was registered that have results uploaded to the registry, by year of record entry. NCAs are ordered by percent of all CTAs reported. Since these trials fall under the oversight of a single NCA there is no ambiguity in where enforcement of EU regulations would sit.

## Discussion

### Summary of Results

There are notable gaps in data quality and availability on the EU Clinical Trials Register. Issues range from missing protocols and results to outdated data on the current status of a given trial. Apparently missing registrations are largely concentrated among a few countries (e.g., France, Romania) while issues with data quality are more widespread. Results availability issues are widespread but concentrated among trials taking place within a single country.

### Strengths and weaknesses

This analysis covers all European trials on the EUCTR as of December 2020 and therefore provides a comprehensive and robust analysis of trends in registration and transparency practices throughout the continent. That said, this is a macro-level examination intended to spot major deviations in registration, data quality, and reporting trends. For instance, it is likely that some Dutch trials have been ongoing with very long follow-up but it is highly unlikely that the current extent of ongoing trials in the Netherlands is accurately reflected in the registered data. Our analysis, however, lacks the precision that would be required to begin to distinguish the extent of mislabeled versus trials with *bona fide* long-term follow-up. Similarly, our investigation into missing CTAs can only act as a proxy for the true extent of the problem as we can only examine this issue for the subset of all trials with tabular results on the EUCTR. Findings in one area also impact the context of other areas. While Romania has the highest percent of single-CTA trials reported it also seems likely that many, if not most, Romanian trials are missing from the registry entirely. There may also be a selection bias in which sponsors that ensure their CTA appears on the public EUCTR website are also more likely to report. Technical limitations of the EUCTR may also impact assessments. The EUCTR only recently implemented reporting procedures for trials that were registered but never occurred and this may not have been acted on by sponsors.^16^ Future work may also seek to understand the local regulatory contexts and detail where and how issues occur. Lastly, we only cover specific aspects of data quality on the EUCTR linked closely to NCA responsibilities and not the overall quality or accuracy of registered information about trial design and conduct.^17,18^

### Findings in Context

We are not aware of any large-scale assessments of data quality and availability on the EUCTR to date. One prior study supports issues with the provision of completion status on the EUCTR compared to ClinicalTrials.gov: 16.2% of trials identified on both registries had a discrepant trial status, the vast majority of which had an “Ongoing” status on the EUCTR but a “Completed” status on ClinicalTrials.gov suggesting lower standard for data accuracy on the EUCTR.^7^ The results of our analysis indicate that issues with incorrect trial statuses continue to appear for a number of high research output countries. Other work has more broadly documented persistent data quality issues across registries, including the EUCTR.^19^

We have encountered many of these data issues in our ongoing EU TrialsTracker work however this analysis represents the first attempt to formally document the problem.^8^ The EU TrialsTracker provides monthly updates on the results status of completed trials on the EUCTR. Without accurate and complete data, public accountability efforts cannot fully operate as intended. As of April 2021, 70.2% of verifiably completed protocols have reported results but many cannot be properly assessed due to data issues with trial completion status and dates. Transparency advocates have been similarly frustrated by these issues in their efforts to improve trial reporting throughout the EU.^20^ Additionally, our EU TrialsTracker work noted large reporting discrepancies between industry and non-industry sponsors, as well as large and small sponsors.^8^ These discrepancies likely account for much of the observed gap in reporting between single-CTA and multi-CTA trials given the frequency of multinational industry-funded trials.^8^

### Policy Implications and Interpretation

Clinical trial registries are a vital source of information to ensure that all clinical trials are reported and that all researchers are transparently accountable to patients, participants, and clinicians. As an ICTRP primary registry, the EUCTR commits to “make all reasonable efforts to ensure that the data registered is complete, meaningful, and accurate.”^21^ EU/EEA countries are a major source of medical research globally and their registration scheme is tied directly to national and EU guidelines and directives meaning nearly even trial on medicines since 2004 should be clearly and publicly documented and reported as part of the standard regulatory process.^5,22–24^ The EUCTR can help plan research priorities, combat reporting biases, and boost evidence synthesis efforts to inform clinical practice but only if records are completed and accurate. France, a major research hub, shows evidence of a large gap in public registrations leaving a pronounced hole in the public European research record. Norway, Poland, and Italy show similar problems and Romania’s issues appear severe. Romania is the 6th most populous country in the EU --it seems impossible that only 239 studies recruited in Romania since 2007. Missing public registration may also complicate publication for researchers who rely on the EUCTR to satisfy ICMJE requirements.^2^ The Netherlands, now home to the EMA, is joined by Spain in having major data issues across their extensive portfolio of trials. National trends, like BfArM posting less research over time compared to their sister Paul Erlich Institute, may also be of interest to local observers.

In response to data quality issues, the UK MHRA noted that records were available in the backend EudraCT system, but further action was required to move them to the public facing EUCTR. Staffing issues were at the root of the UK delays and were swiftly managed after they were brought to the attention of a Parliamentary committee.^14,15^ This may be informative to other NCAs. In the best case scenario the proper paperwork and other regulatory materials are held by NCAs but have not yet been acted upon. Addressing issues could be rectified through concerted efforts to improve record-keeping and data-entry tasks related to the trial database. It is also important to understand to what extent issues originate with sponsors. The EMA has conducted some proactive outreach to remind sponsors of their responsibility to report but NCAs would be expected to have more direct and frequent engagement with local sponsors to rectify specific issues.^25^ The Austrian NCA conducted outreach to sponsors directly about their reporting responsibilities and has seen subsequent increases in results submissions.^26^ It is also nonsensical for sponsors to have trials with mismatching information within their registrations. These make entries on the registry difficult to search, understand, and analyse for users. Flexibility in working with sponsors outside rigid bureaucratic rules, especially in rectifying data from very old trials, may be warranted. Proactive outreach and education from NCAs and the EMA about improving data quality and results availability may also be necessary to promote improvement at scale.

Major gaps and shortcomings in such a vital database should at the very least be transparently documented. Ideally, regular public audits by the EMA would identify these issues and address them at the source. If regulatory processes in these countries are operating as intended, data on fundamental aspects of a trial such as when it recieved the proper approvals or when it completed should be readily available and flow unobstructed to the public register. We hope the EMA will closely examine what has become of this missing data and support efforts to improve the reliability and validity of the public EUCTR dataset and transparently audit NCA-level progress in fulfilling their responsibilities. The Heads of Medicines Agencies (HMA) organisation, a network of EU NCA leadership, may be an effective partner for coordinating improvement and sharing best practices between NCAs. The HMA has recently announced plans to further encourage reporting to the EUCTR in response to external pressure.^27,28^ While the UK is no longer a member of the EU or the HMA, their high performance across the investigated areas suggests the the MHRA may have key learnings to share with the European regulatory community. Hopefully their current political distance will not act as a barrier to this knowledge exchange.

A new EU trial portal is set to launch in January 2022. However, the EUCTR should not be neglected as an important source of clinical trial information. The corpus of registered trials from 2004 through the 2023 phase-out of new registrations on the EUCTR should contain evidence on many treatments in wide use today.^29^ While data management in the new portal will change, NCAS will still play an important oversight role.^30^ Individual countries are also empowered to sanction non-compliant sponsors.^23^ Key learnings from the current clinical trial regulations should inform staffing needs and internal processes moving forward. NCAs should therefore ensure they have adequate resourcing and plans to monitor data quality and reporting that falls under within their jurisdiction.

## Conclusion

There are persistent and notable gaps in the quality and completeness of trial data on the EUCTR. The public dataset appears to be missing registrations with over half of all checked trials missing CTAs for France, Romania, and Norway. Additional major European clinical research hubs like Spain and The Netherlands have substantial issues with data quality and results availability. The processes that guide the collection and dissemination of this data are embedded in a clear regulatory structure so their apparent failure is concerning. Users of the EUCTR, including researchers, governments, clinical guideline developers, and the public would benefit from a more complete and accurate accounting of the European research environment via the official EU registry and steps should be taken to ensure NCA-level issues are proactively and transparently identified, documented, and addressed.

## Supporting information

STROBE Checklist

## Acknowledgements

The authors would like to thank our colleagues at the DataLab for their ongoing support of the TrialsTracker project and feedback on this manuscript.

## Author Contributions

NJD collected, cleaned, and analysed, and wrote the first draft of the manuscript with supervision from BG.

