## Supplementary Information for "Trends and Variation in Data Quality on the EU Clinical Trials Register: A Cross-Sectional Study"

### Supplementary Information: DeVito & Goldacre

#### Supplemental Figure 1

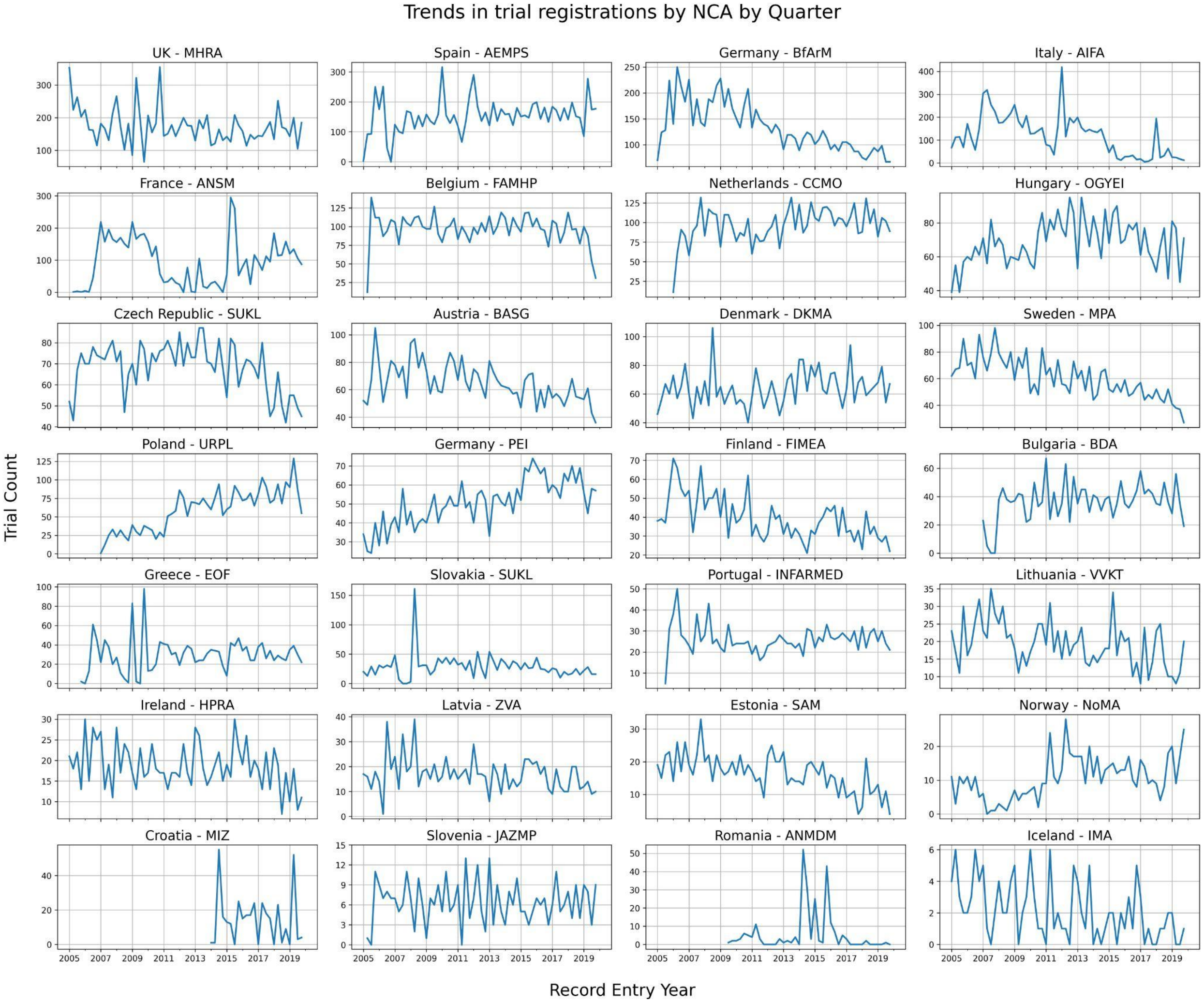

### Supplemental Figure 2

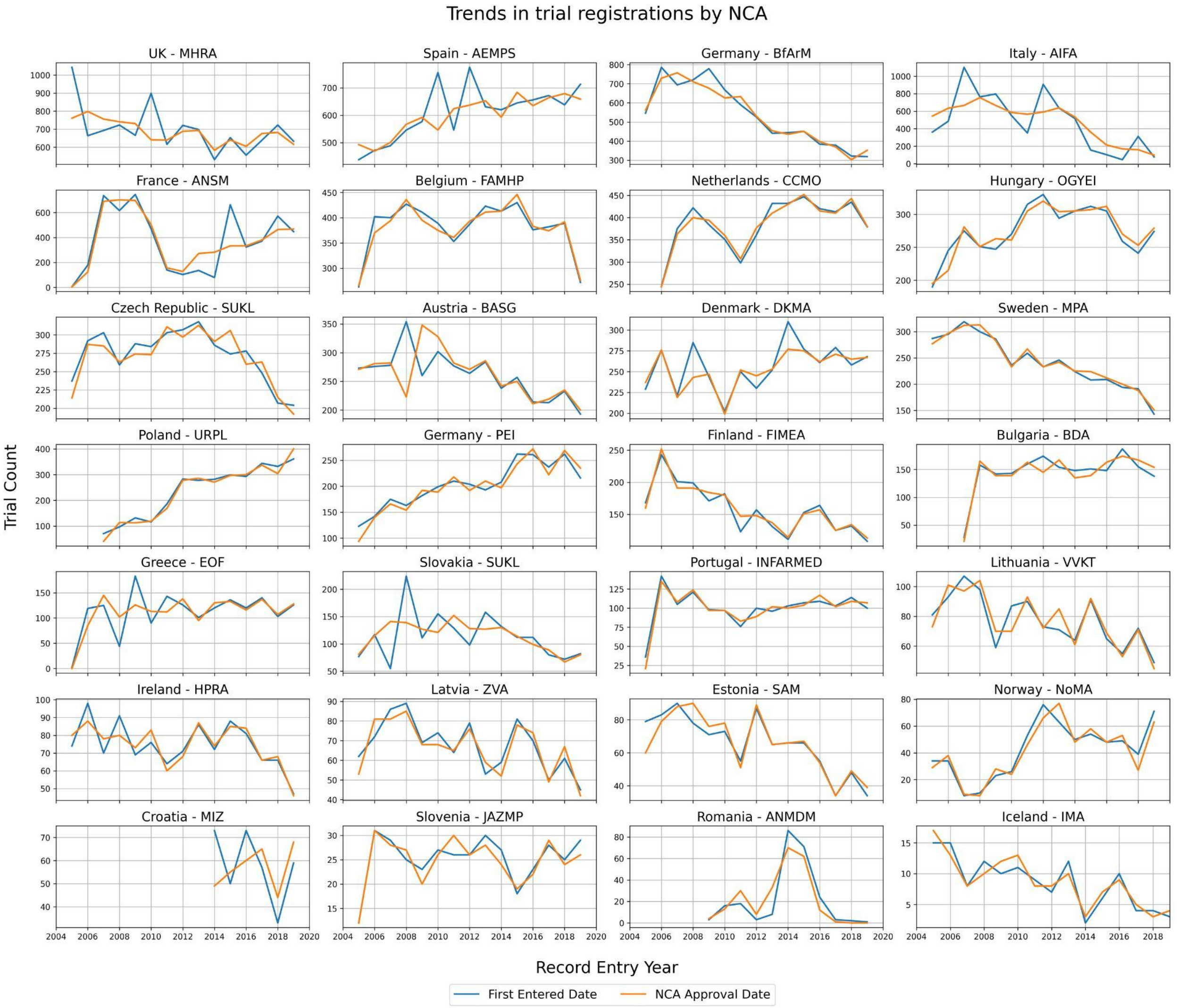

#### Supplemental Figure 3

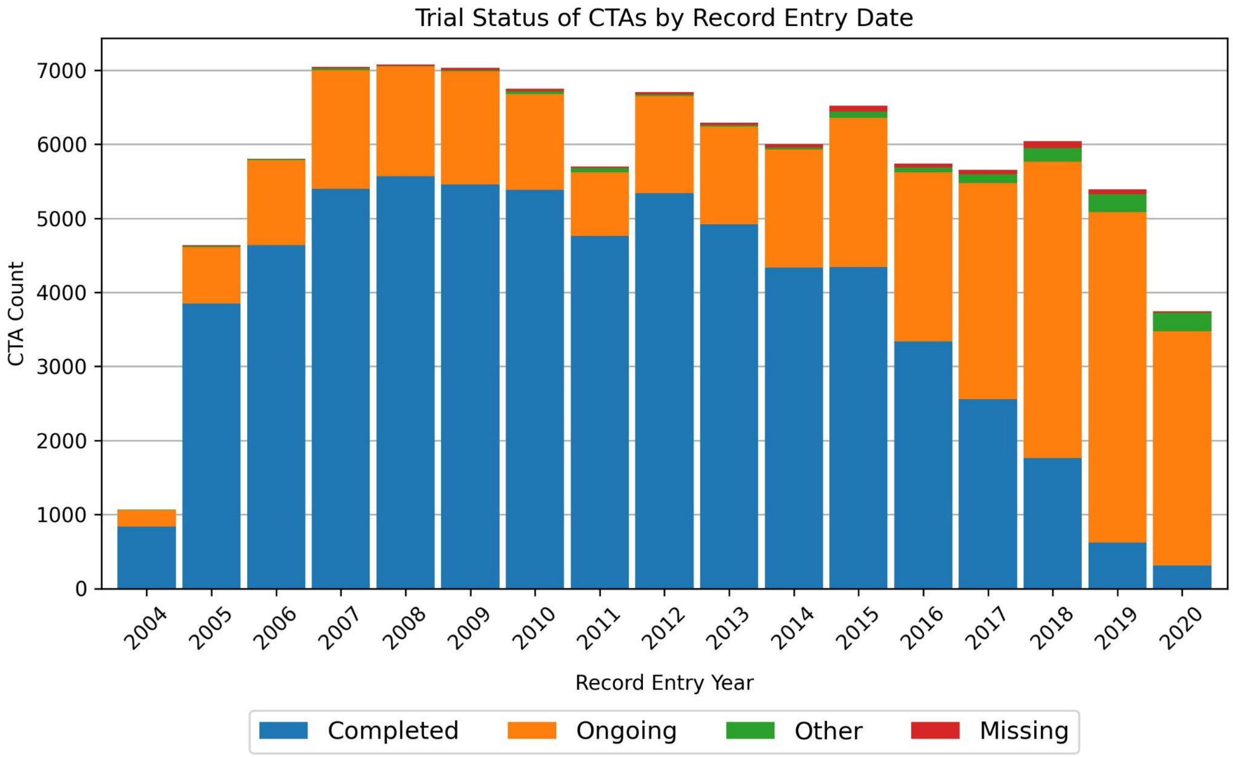


#### Supplemental Figure 4

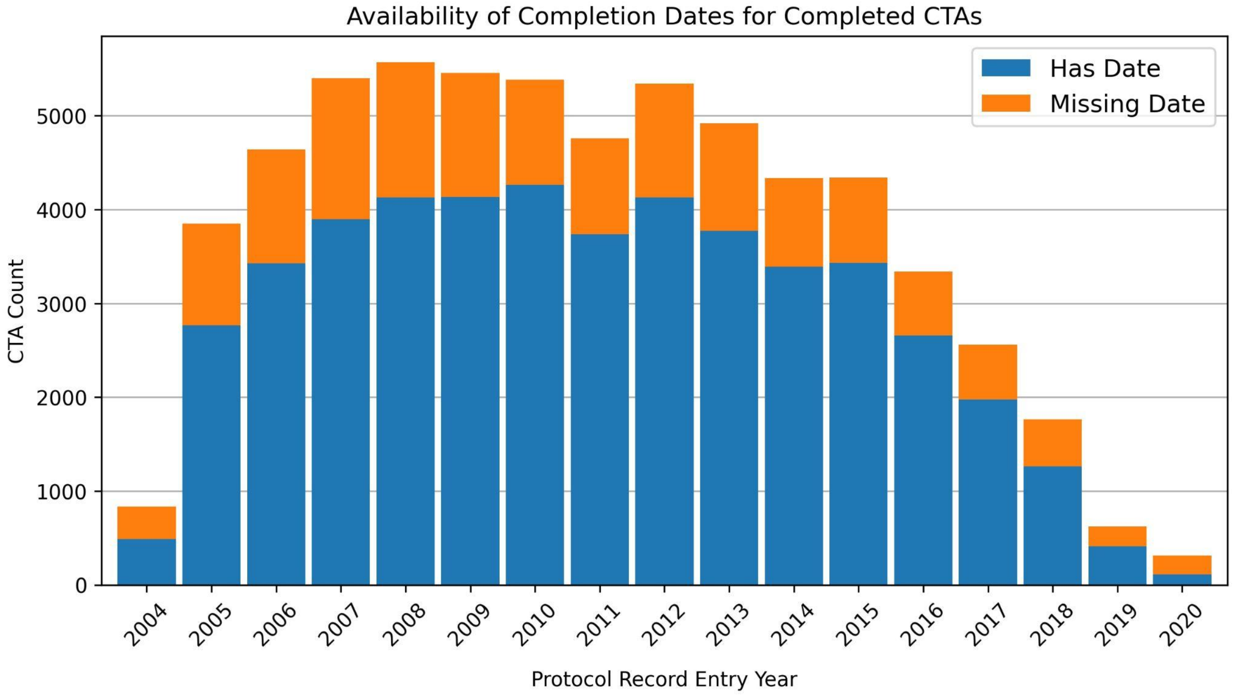
